# Current status and expectations of medical interns in the advanced phase of the DES (specialized medical studies) in medical gynecology regarding medical theses

**DOI:** 10.1101/2025.01.23.24319156

**Authors:** Aurelie Poisson, Floriane Jochum, Paul Gougis, Raphaelle Savarino, Florence Coussy, Charlotte Sonigo, Enora Laas, Damien Roux, Fabien Reyal, Christine Rousset-Jablonski, Anne-Sophie Hamy

## Abstract

**Introduction:** In France, Writing and defending a medical thesis are mandatory components of medical education curriculum to obtain the degree of Doctor of Medicine. The expectations of medical interns in medical gynecology regarding their medical thesis have not been evaluated.

**Methods:** A 10-question form was distributed during the national teaching of the advanced phase of the specialized medical studies (DES) in medical gynecology (corresponding to the 2^nd^ and 3^rd^ year of internship). Responses were collected using the Wooclap* software.

**Results:** A total of 131 interns participated in the survey out of 170 interns solicited. Approximately two-thirds of the surveyed students had a medical thesis topic at the time of the survey. Among students with a thesis topic, thesis supervisors were most often university practitioners (MCU-PH/PH-U/PU-PH, 37.4%), followed by hospital practitioners (22.8%). The quality of supervision by the thesis supervisor was rated at 6.9/10, and the quality of supervision by the university was rated at 3.3/10. Overall, 53.4% of students wished to leverage their work through scientific publication. Nearly all students (94%) wished for guidance on at least one of the following topics: writing scientific articles (83%), basic statistics (83%), basic data collection (75%), training in the use of Pubmed (68%), Excel (65%), Zotero (63%), project management (53%), Word (15%).

**Conclusion:** Support for medical interns in medical gynecology for their medical thesis could be improved, particularly through modules on scientific publication, bibliography, and office automation.

## Introduction

The Specialized Studies Diploma (DES) in medical gynecology was created in 2003 (Serfaty, 2000) (Agopiantz et al., 2013, 2013). It currently includes a specialized 4-year training, which can be complemented by a transversal specialized training year (FST) in the fields of oncology, reproductive medicine and biology - andrology, or medical/pharmacological therapeutics. In addition to hospital internships, a volume of two half-days per week of theoretical teaching is planned, including one half-day of autonomy. The theoretical teaching is provided in the form of face-to-face lectures, clinical cases on the specialty college’s digital platform, and e-learning sessions validated by regional coordinators.

In 2015, an evaluation of the theoretical teaching of the DES had been conducted with 145 students. Despite a rather satisfactory evaluation, avenues for improvement had emerged, including the creation of specific teaching modules, situational exercises, and practical work (Agopiantz et al., 2015).

In France, the award of the title of Doctor of Medicine is conditioned by the writing of a medical thesis manuscript, publicly defended during a defense in front of a jury. Although encouraged, the publication of the medical thesis is not mandatory. To our knowledge, no data have been published on the theses carried out by interns in medical gynecology since its creation in 2023, and the expectations of medical interns in medical gynecology regarding their medical thesis have not been evaluated. The aim of this study is to assess the current status of medical interns in medical gynecology in the advanced phase regarding the supervision of their medical thesis and their desire for training and supervision.

## Materials and methods

### Survey design and sampled population

We conducted a survey on March 9, 2023, during the national teaching of senology of the DES in medical gynecology. This teaching was carried out remotely, synchronously, via the Teams* platform (Microsoft), and concerned interns in the advanced phase. Interns were invited to respond to an interactive questionnaire created via the Wooclap* platform.

### Questionnaire

The questionnaire (Table 1) consisted of 10 questions: 6 single-choice questions, 2 multiple-choice questions, while 2 questions used a 0 to 10 rating scale. Completing the questionnaire in its entirety required less than 10 minutes, and its completion was anonymized.

**Table 1:** Questionnaire. Questions 4 and 10 were multiple choice. Questions 7 and 9 used a rating scale from 0 to 10. The remaining 6 questions were single choice.

| Qx. | Question | Answers |
| --- | --- | --- |
| Q1. | What is your original subdivision? |  |
| Q2. | Do you have a medical thesis topic? | 1. Yes. / 2. No. / 3. I don't know. |
| Q3. | Have you already abandoned one or more thesis topics? | 1. Yes. / 2. No. / 3. I don't know. |
| Q4. | If yes, for what reasons? | 1. Poor initial definition of the topic<br>2. Lack of interest in the proposed topic<br>3. Workload<br>4. Relational conflict<br>5. Financial feasibility<br>6. Negative results<br>7. Lack of guidance |
| Q5. | What is the position of the main person supervising you? | 1. University hospital practitioner (MCU/PHU/PUPH)<br>2. Hospital practitioner (PH)<br>3. Clinical assistant (CCA)<br>4. Assistant<br>5. Other<br>6. Not applicable |
| Q6. | Is this person part of the department where you would like to do your post-internship? | 1. Yes. / 2. No. / 3. Not applicable. |
| Q7. | How do you rate the quality of supervision by your thesis supervisor? | Rating scale from 0 to 10. |
| Q8. | Do you wish to enhance your thesis through scientific publication? | 1. Yes. / 2. No. / 3. I don't know. |
| Q9. | How do you rate the quality of supervision for your thesis by your original UNIVERSITY? | Rating scale from 0 to 10. |
| Q10. | Would you like guidance on the following topics? | 1. Word<br>2. Excel<br>3. Zotero<br>4. Pubmed<br>5. Database collection<br>6. Project management<br>7. Basic statistics<br>8. Scientific article writing |

### Ethical oversight

The local institutional review Board of institute Curie reviewed this project on 16/12/2024 under the number DATA240346 and confirmed that it does not require formal ethical oversight, as it qualifies for exemption under the applicable ethical guidelines for non-invasive and anonymized research. The committee’s decision was as follows: Ethical oversight was waived for this study.

### Statistical analysis

Subjective quantitative variables were collected using a 0 to 10 scale. Medians (interquartile ranges, IQR) and frequencies (percentage) were calculated to respectively describe continuous and categorical variables. Proportions were compared using chi-square tests. Data were analyzed using R software, version 3.2.1.

## Results

### Population characteristics

A total of 131 interns participated in the survey (out of 170 interns enrolled in the advanced phase), with 106 completing the questionnaire in its entirety. The three most represented subdivisions were the Paris subdivision, followed by Lille and Bordeaux (Figure 1). Most interns had a medical thesis topic (n=66, 62.3%), while about one-third of surveyed students (n=34) did not have a medical thesis topic (do not know 5.6%).

**Figure 1:**
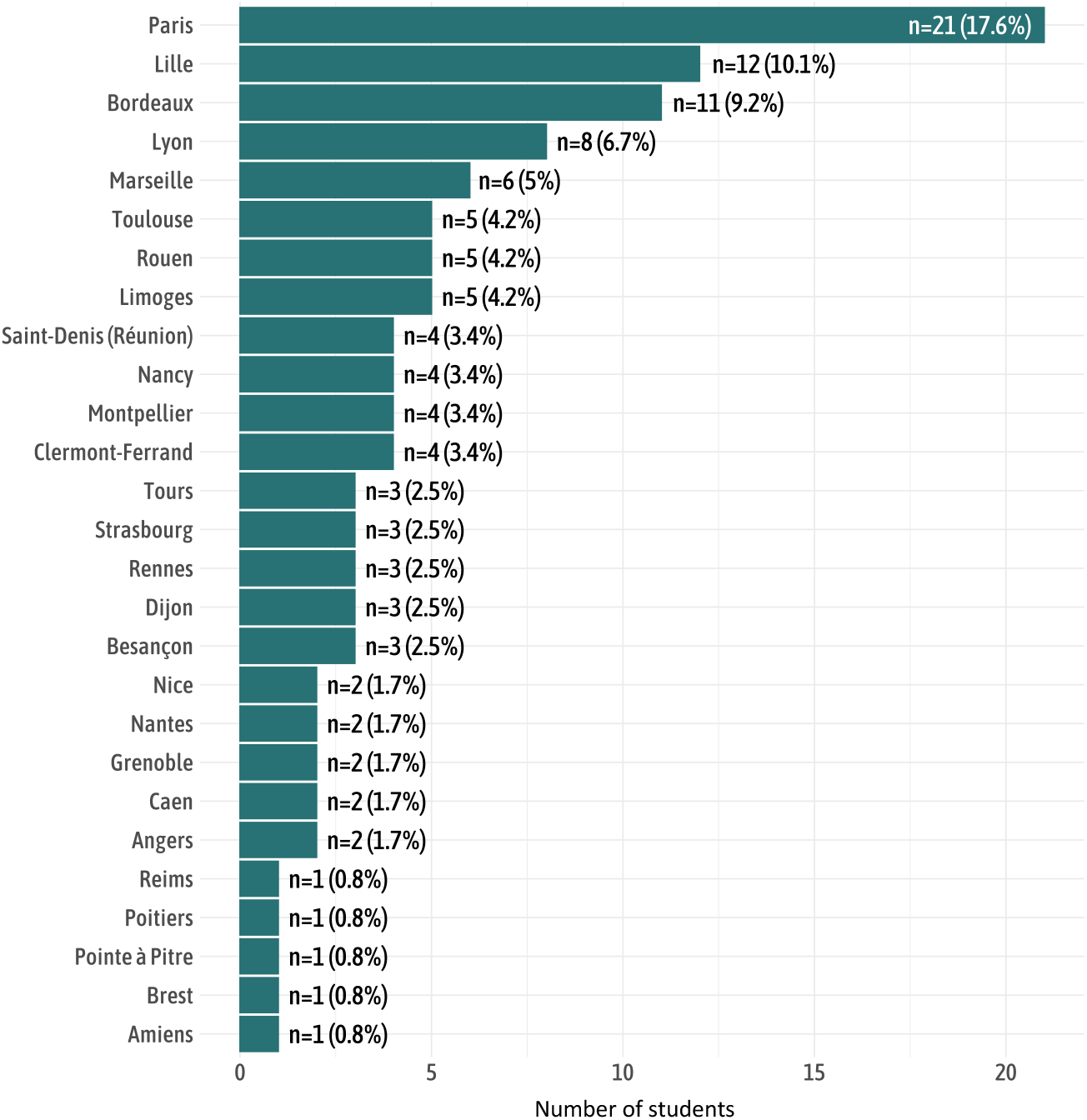
Distribution of subdivisions of medical gynecology interns who responded to the questionnaire.

### Medical thesis topic abandonment

Twenty-nine students (27.6%) had already abandoned one or more thesis topics, with the two main reasons cited being the initial poor definition of the topic (37.0%) and lack of supervision (33.3%) (Figure 2).

**Figure 2:**
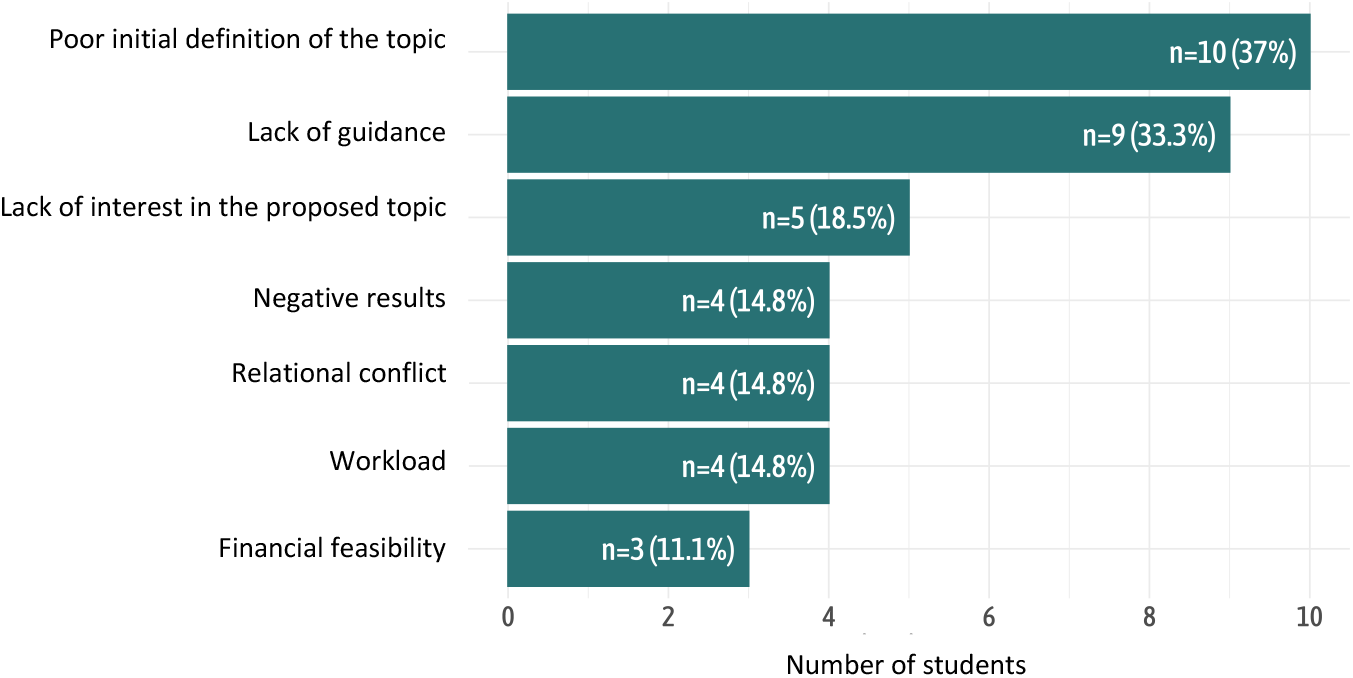
Distribution of reasons for abandonment among interns who changed their thesis topic.

### Supervision of the medical thesis

Among students with a thesis topic, thesis supervisors were most often university practitioners (MCU/PHU/PUPH, 37.4%), followed by hospital practitioners (22.8%), while supervision by CCA and AHU was marginal.

Most supervisors were not part of the department in which the student wished to complete their post-internship (n = 54, or 66% in total), while 28 supervisors (34%) were part of it.

### Evaluation of the quality of supervision for the medical thesis

The quality of supervision by the thesis advisor was rated at 6.9/10, while the quality of supervision by the university was rated at 3.3/10 (Figure 3). Overall, 53.4% of students wanted to promote their thesis through a scientific publication, 24.3% were unsure, and 22.3% did not wish to do so.

**Figure 3:**
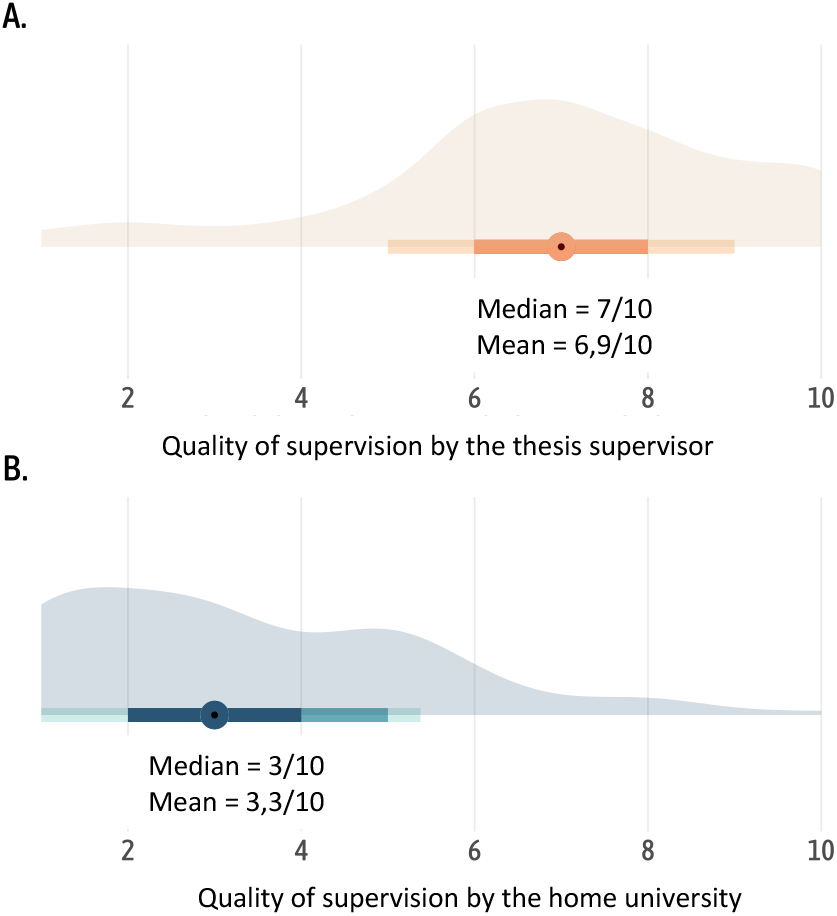
A. Mean and median of the quality of supervision by the thesis supervisor. B. Mean and median of the quality of supervision by the home university.

## Interns’ reported needs regarding medical thesis supervision

Nearly all students (114/131, 87.0%) desired supervision on at least one of the following topics: Scientific article writing (83%), basic statistics (83%), database collection (75%), PubMed training (68%), Excel (65%), Zotero (63%), project management (53%), Word (15%) (Figure 4).

**Figure 4:**
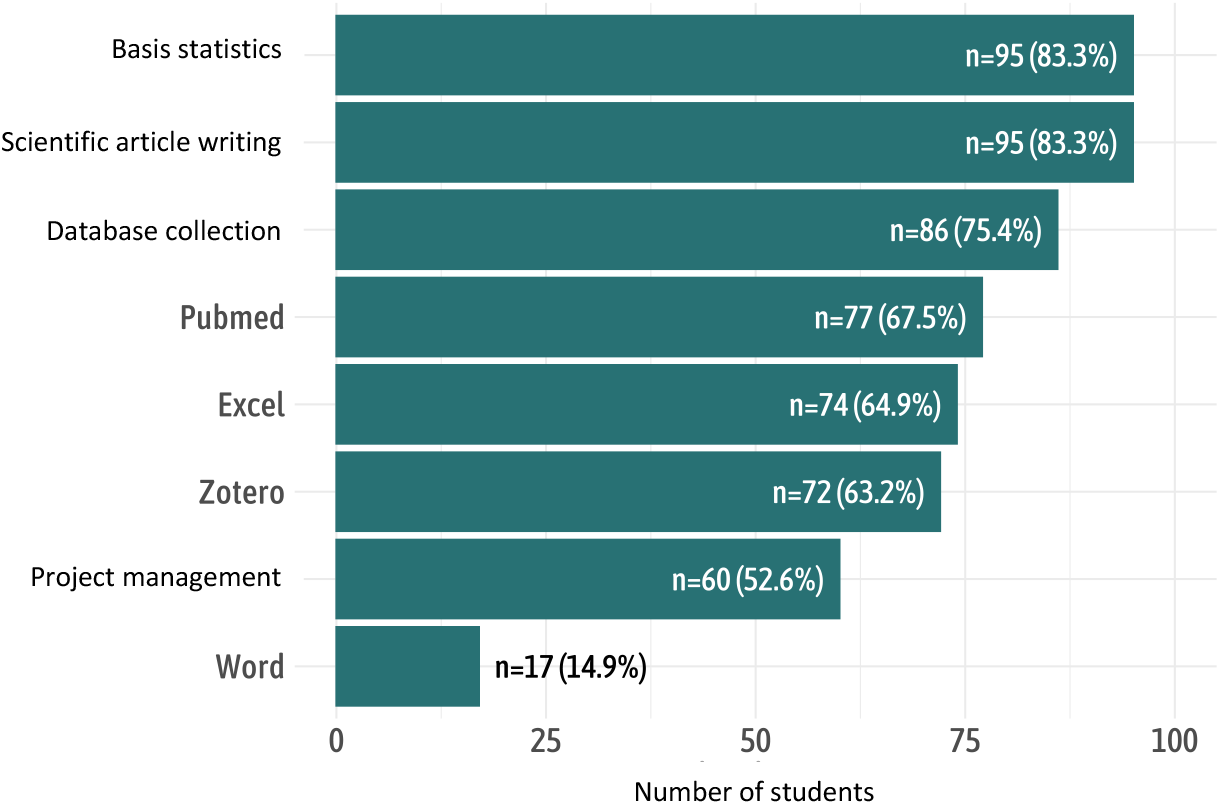
Proportion of interns wishing for additional training in one of the following areas.

## Discussion

Our survey enabled us to analyze the expectations of senior-phase medical gynecology interns regarding their medical theses. This study provides several important findings.

Firstly, most interns already had a thesis topic during their advanced phase, in line with the reform of the third cycle of medical studies, which mandates the completion of the medical thesis before the start of the fourth year. Students were primarily supervised by senior university practitioners, highlighting the active involvement of academic staff in overseeing medical theses. Only 34% of supervisors were part of the department where the intern intended to pursue their post-internship, suggesting a relative independence between the intern and their supervisor concerning their future professional pathway.

While the students’ rating of the direct supervision by their thesis advisor appeared satisfactory, university-level supervision seems to require improvement. To our knowledge, few programs specifically aimed at supporting third-cycle students in completing their medical thesis are offered across various faculties. In Germany, a program was initiated by a consortium of universities, which allows: (i) identifying students’ needs; (ii) establishing a research support program; and (iii) implementing a process of continuous improvement that considers developments in the literature and student satisfaction. Areas requiring support included principles of good scientific practice, literature searches, reference organization, the structure and organization of a thesis, formatting word processing documents, clinical epidemiology, and data management (Paulitsch et al., 2016; Sennekamp et al., 2016). Implementing similar programs in France would seem worthwhile.

Nearly one in five interns reported previously quitting a thesis topic, mainly due to poor initial definition of the topic or lack of supervision. In a survey conducted by a young oncology cooperative group (Dig’In for GERCOR), 27.4% of 143 oncology interns reported having already abandoned a medical thesis topic, with the main reason also being poor initial topic definition. In this regard, university-level training for medical thesis supervisors could also be considered to reduce the proportion of abandoned thesis topics.

Finally, most interns appeared interested in promoting their thesis through a scientific publication. In France, few studies have evaluated the publication rates of medical theses. Analysis of data from the Faculty of Medicine in Lille (2001-2007) found a publication rate of 11.3%. According to other studies, the publication rates for medical theses were approximately 6% for general medicine theses (Baufreton et al., 2012; Carpentier et al., 2012) and 20% for theses in other specialties (Fabre, 2015), with an estimated overall average of 17% across all specialties (Salmi et al., 2001). In radiology, the publication rate for interns graduating in 2009-2010 was 35.3% (Chassagnon et al., 2016), while the rate for theses completed by medical oncology interns between 2010 and 2015 was 29.7% (de Nonneville et al., 2022). In medical gynecology, no data exist on the publication rate of medical theses, but our study suggests a willingness among most current cohorts of medical gynecology interns to publish their work.

In conclusion, medical gynecology interns appear eager for structured supervision modules to help them complete their medical theses. Such programs could facilitate the promotion of their work through scientific publications. A pilot program will soon enter a testing phase at Paris Cité University.

## Data Availability

All data produced in the present work are contained in the manuscript

